# Precautionary breaks: planned, limited duration circuit breaks to control the prevalence of COVID-19

**DOI:** 10.1101/2020.10.13.20211813

**Authors:** Matt J. Keeling, Glen Guyver-Fletcher, Alex Holmes, Louise Dyson, Michael J. Tildesley, Edward M. Hill, Graham F. Medley

**Author notes:** These authors contributed equally to this work (Louise Dyson, Michael J. Tildesley, Edward M. Hill).

## Abstract

The COVID-19 pandemic in the UK has been characterised by periods of exponential growth and decline, as different non-pharmaceutical interventions (NPIs) are brought into play. During the early uncontrolled phase of the outbreak (early March 2020) there was a period of prolonged exponential growth with epidemiological observations such as hospitalisation doubling every 3-4 days (growth rate *r* ≈ 0.2). The enforcement of strict lockdown measures led to a noticeable decline in all epidemic quantities (*r* ≈ −0.06) that slowed during the summer as control measures were relaxed (*r* ≈ −0.02). Since August, infections, hospitalisations and deaths have been rising (precise estimation of the current growth rate is difficult due to extreme regional heterogeneity and temporal lags between the different epidemiological observations) and various NPIs have been applied locally throughout the UK in response.

Controlling any rise in infection is a compromise between public health and societal costs, with more stringent NPIs reducing cases but damaging the economy and restricting freedoms. Currently, NPI imposition is made in response to the epidemiological state, are of indefinite length and are often imposed at short notice, greatly increasing the negative impact. An alternative approach is to consider planned, limited duration periods of strict NPIs aiming to purposefully reduce prevalence before such emergency NPIs are required. These “precautionary breaks” may offer a means of keeping control of the epidemic, while their fixed duration and the forewarning may limit their society impact. Here, using simple analysis and age-structured models matched to the unfolding UK epidemic, we investigate the action of precautionary breaks. In particular we consider their impact on the prevalence of infection, as well as the total number of predicted hospitalisations and deaths. We find that precautionary breaks provide the biggest gains when the growth rate is low, but offer a much needed brake on increasing infection when the growth rate is higher, potentially allowing other measures (such as contact tracing) to regain control.

## 1 Introduction

The novel coronavirus virus strain that arose in the Wuhan city in China in late 2019 has had a dramatic effect on the lives of people worldwide. By 1st October, we had passed the grim milestone of 1 million deaths worldwide and over 30 million cases [1]. In the absence of disease specific treatments or prophylactic measures, most countries have adopted social distancing measures (reducing the number of potentially risky contacts) as a means of control [2]. The optimal level of non-pharmaceutical interventions (NPIs) requires a careful balance between public health and economic needs, from which it could be argued that the optimal strategy is one that minimises economic disruption while still preventing exponential growth of infection [3].

In the UK, the first cases of COVID-19 were reported on 31st January 2020 in the city of York. Cases continued to be reported sporadically throughout February and by the end of the month guidance was issued stating that travellers from the high-risk epidemic hotspots of Hubei province in China, Iran and South Korea should self-isolate upon arrival in the UK. By mid-March, as the number of cases clearly began to rise, there was advice against all non-essential travel and, over the coming days, several social-distancing measures were introduced including the closing of schools, non-essential shops, pubs and restaurants. This culminated in the introduction of a UK “lockdown”, announced on the evening of 23rd March, whereby the public were asked to remain at home with four exceptions: shopping for essentials; any medical emergency; for one form of exercise per day; and to travel to work if absolutely necessary. By mid-April, these stringent mitigation strategies began to have a noticeable effect, as the number of confirmed cases, hospitalisations and deaths as a result of the disease began to decline[4, 5]. As the number of daily confirmed cases continued to decline during April, May and into June[6], measures to ease lockdown restrictions began, with the re-opening of some non-essential businesses and allowing small groups of individuals from different households to meet up outdoors, whilst maintaining social distancing. This was followed by gradually re-opening primary schools in England from 1st June and all non-essential retail outlets from 15th June.

The dynamics throughout the summer of 2020 were characterised by the emergence of hot-spots of persistent infection and higher than average growth rates, with such areas facing additional local controls. Notably, Leicester and the Greater Manchester conurbation were identified as regions where the number of identified cases per hundred thousand had increased to high levels, while the infection in many other areas was still in decline. By mid to late August, there was some evidence that the national underlying growth rate had become positive [6], and by the end of September exponential growth had returned in almost all regions [7, 8]. Like many countries, the UK is now attempting to curtail growth by introducing stricter controls, but “lockdown” is very disruptive to multiple elements of society [9, 10], especially given that restrictions are largely unpredictable to the local populous and businesses.

One potential mechanism of regaining control is to introduce a short period of intense measures to substantially reduce cases — this has been dubbed a ‘circuit breaker’ although “precautionary break” is a more apposite name. It is hoped that by driving infections to a sufficiently low level, other measures such as test-trace-and-isolate will have greater capacity to prevent the spread of infection [11]. In common with other resource limited controls [12], we expect test-trace-and-isolate to be most effective when the level of infection is relatively low [13]. Additionally, a short lockdown period would limit the economic costs of such a measure.

In this paper, we consider the application of breaks to the UK, aligning a two-week period of intense control to school half-terms in order to minimise educational disruption. We utilise both a simple illustrative analysis and an age-structured model fitted to the UK data, to investigate the likely impact of a break on the trajectory of infection and the subsequent numbers of hospitalised cases and deaths.

## 2 Simple Analysis

Under the simplifying assumption that without additional control measures cases will increase exponentially at rate *r*(> 0), but during the period of the precautionary break cases will decrease at a rate −*s*(< 0), we can develop a simple model for the dynamics:

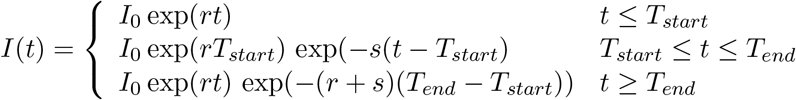

where *T*_*start*_ and *T*_*end*_ define the start and end dates of the breaks. From this analysis, we observed two key effects of a short-term lockdown. Firstly, it causes a relative reduction in the level of infection by a factor *B* = 1 − exp(−(*r* + *s*)(*T*_*end*_ − *T*_*start*_)), compared to not having the precautionary break. In principle, this should translate into a similar reduction in the daily number of hospitalisations and deaths - the two epidemiological measures of most concern. We can also conceptualise this reduction as taking the epidemic back in time to when there were fewer cases; this temporal reset, *T*_*R*_, is given by:

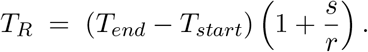

Taken together these two metrics *B* and *T*_*R*_ imply that such breaks, unsurprisingly, have the greatest impact when they are long in duration and lead to a rapid decline in cases (*s* large). The impact of the uncontrolled growth rate, *r*, is more ambiguous. When *r* is large the impact of the break on the level of infection is more pronounced as it prevents a period of high exponential growth. In contrast, large *r* reduces the size of the temporal reset as it does not take many days of exponential growth to cancel the decline expected during the break.

## 3 Age-structured model description

We now wish to increase the realism of the modelling framework to more accurately capture the impact of such breaks on the number of hospitalisations and deaths as a result of infection, which requires an age-structured model matched to the wealth of epidemiological data [14, 15]. Even though we have dramatically increased the complexity of the model, the two simple metrics (*B* and *T*_*R*_) remain of key interest.

The age-structured model and its matching to the UK data has been described in detail elsewhere [14, 15]. Here, for completeness, we provide a basic review of the main salient points. The model is an ODE formulation based around SEIR-type dynamics, and incorporating five-year age classes. The infection dynamics are modified to account for symptomatic and asymptomatic infection (dependent on age), age-dependent susceptibility and reduced transmission from asymptomatic infections. The model also includes additional structure to account for quarantining of identified cases and isolation of their household.

The model is calibrated against different sources of epidemiological data including hospitalisations, admission to ICU, deaths, age-structured serology from blood donors and the results of pillar 2 test-bing [15]. This matching is done separately for the seven NHS regions in England (East of England, London, Midlands, North East & Yorkshire, North West, South East and South West) and for the three devolved nations (Northern Ireland, Scotland and Wales); although some parameters (such as the relative transmission rate from asymptomatic infections) are universal. In particular, we highlight that the historical and changing pattern of regulations, restrictions and behaviour in each region is captured by a weekly rescaling of the transmission matrix.

We consider two distinct forms of simulation. In the first, we tightly constrain the dynamics (by rescaling the transmission matrix) to have a given growth rate *r* from 1st September, with the growth rate being estimated by an eigenvector approach. During the 2-week break (24th October - 7th November) we again rescale the transmission matrix to achieved the desired rate of decay −*s*, before returning to the pre-break matrix. All simulations are performed using a single parameter set.

In the second set of simulations, we allow more regional heterogeneity by fully exploring the posterior parameter estimates. We replicate a range of NPIs by multiple rescalings of the transmission matrix, and set the behaviour in the break to coincide with regional estimates of transmission at the height of the March/April lockdown.

## 4 Results

To illustrate the simple analytical behaviour in more detail, we pick a definitive case (Figure 1): a two-week break from 24th October -7th November to coincide with school half-terms (thereby minimising educational disruption). We consider the relative reduction *B* and the temporal reset *T*_*R*_ across a range of positive growth rates *r*, from 0.01 to 0.22, corresponding to the upper bound of the growth in the March. We consider five different intensities of break (*s* = 0, 0.02, 0.045, 0.06 and 0.1) with the first four aligning with different periods of control in the UK (August, June, May, April), and the latter corresponding to the observed decline in continental Europe where the lockdown restrictions were more intense. We find that, as shown earlier analytically, the greatest effects are found for stricter controls during the precautionary break; lower intrinsic growth (*r*) outside of the break leads to a longer temporal reset, whereas a higher growth rate is associated with a bigger relative decline in infection.

**Fig. 1:**
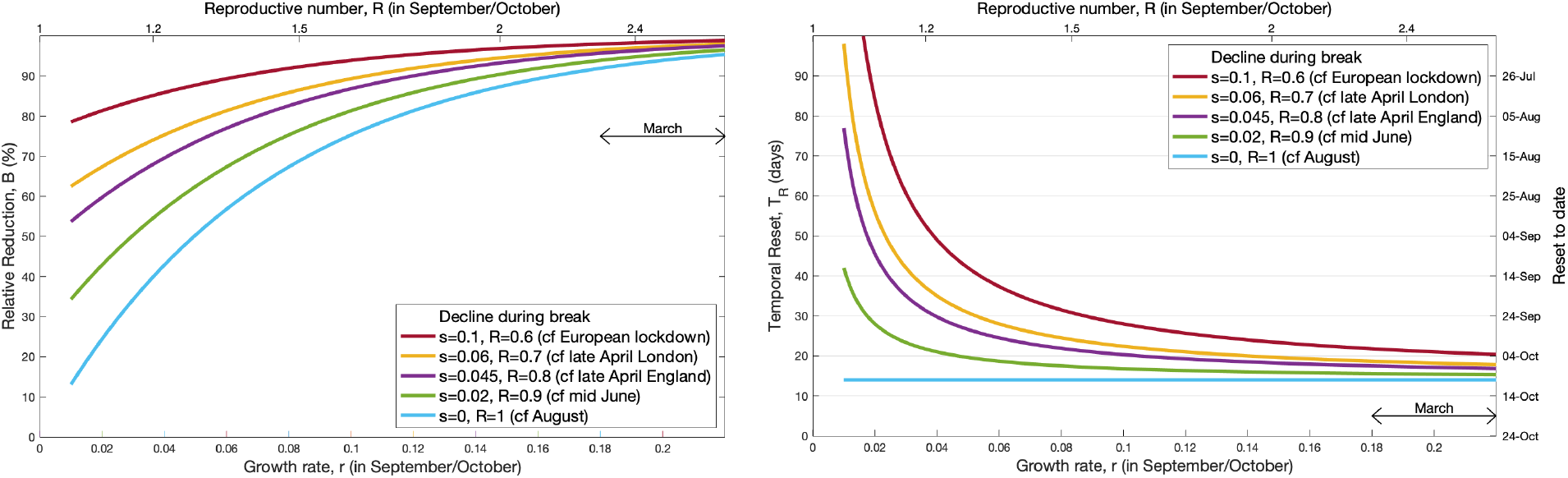
Impact of a 2-week precautionary break during half-term on the infection dynamics. For a range of assumed growth rates in September and October, we illustrate both the reduction in infection (*B*) and the temporal reset (*T*_*R*_) of a break starting on 24th October. Each line corresponds to a different strength of control during the two week break, which is linked to observed epidemic declines.

For the age-structured model, we initially focus on the temporal dynamics over the period from August to the end of the year (Figure 2). We chose four rates of decline during the break corresponding to different rates observed in the UK under different lockdown measures (using the same colours as in Figure 1). For comparison we also show the dynamics without a 2-week break, but including the standard 1-week half term break for school children (black dashed line). We contrast the behaviour at four different intrinsic growth rates for the period 1st September to 24th October (*r* ≈ 0.02, *r* ≈ 0.03, *r* ≈ 0.05 and *r* ≈ 0.07, columns of Figure 2). Given the extra realism of the age-structured model, we project not just the number of infections (top row), but the number of daily hospitalisations and deaths (middle and bottom row respectively). We also give the total number of infections, hospitalisations and deaths between 1st October and 1st January as numerical values.

**Fig. 2:**
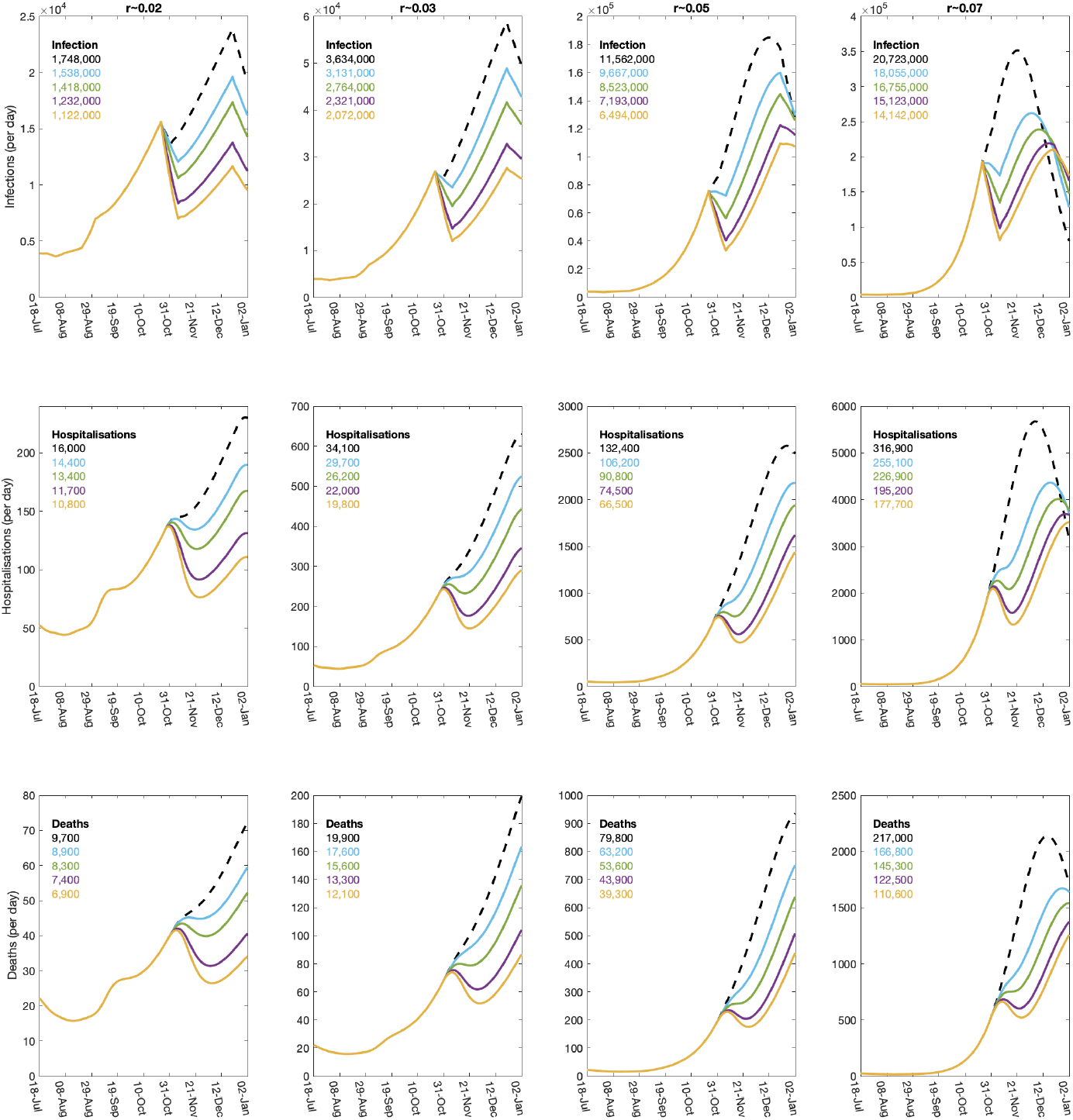
Impact of a 2-week precautionary break during half-term on the dynamics of the full age-structured model. The columns are for four different underlying epidemiological growth rates (*r* ≈ 0.02, 0.03, 0.05 and 0.07, while within each panel the colours correspond to different rates of decline during the half-term break (dashed black, no-control *s* = −*r*; blue *s* = 0; green *s* = 0.02; purple *s* = 0.045; gold *s* = 0.06). The top row shows the number of new infections in the UK, which experiences the most immediate impact of the break; the middle row shows the number of daily hospitalised infections; while the lower row shows the daily mortality.

These results reflect the findings from the simple analytical framework, but provide greater clarity on the impact of these breaks on the key epidemiological quantities of hospitalisations and deaths. Both of these lag behind the changes in infection, and the drop in both quantities after the half-term break is far smoother than is observed for infection. For all three measures, it is clear that the temporal reset increases with the strength of controls during the break, but decreases with the growth rate; the reset is also longer and more clearly defined for infection than for the lagged measures of hospitalisations and deaths. Looking at the medium term impact of the break (as measured by the total number of deaths between 1st October and 1st January), we observe that for low growth rates the strongest levels of NPI during the break reduces deaths by approximately 29%. In contrast, when the growth is high the reduction is approximately 49%.

The results of Figure 2 artificially constrain the dynamics to achieve fixed rates of growth and decline. To consider the potential dynamics in more detail we chose 1000 samples from the posterior distribution [15], for each region of the UK, and vary the level of NPIs to achieve a range of growth rates (*r*). A two-week break is enacted on 24th October, with the rate of decline (−*s*) determined by the maximal level of NPI over the full epidemic to date (as estimated in the posterior parameter distribution); hence introducing far more variability into the process. We again looked at the temporal reset (*T*_*R*_) defined as the time difference between the end of the break and the previous time when a similar level of infection occurred.

The results of these replicates that include natural parameter and control uncertainty show remarkable agreement with the simple analysis (depicted by the red line, Figure 3a). The exception is for the very highest levels of considered growth rates (*r* > 0.07); allowing this level of increase since early September leads to depletion of the susceptible population, which brings a greater temporal reset than expected. However, allowing a growth rate of *r* > 0.07 (a doubling time of less than 10 days) for a long period is not a viable public health option. We find a similar pattern when examining the relative reduction in deaths; in keeping with simple theory there is a rise in the reduction that can be obtained as the growth rate increases, but this again suffers from the non-linear effects of susceptible depletion above *r* = 0.07.

**Fig. 3:**
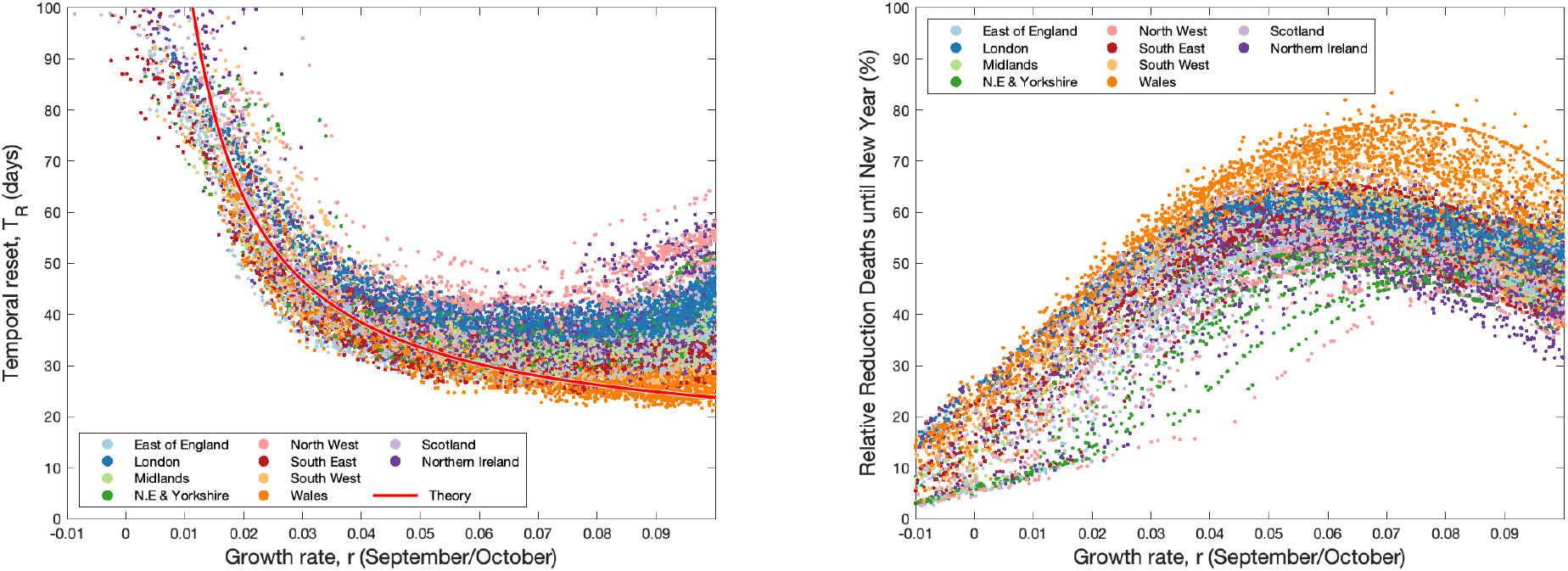
Impact of a 2-week precautionary break during half-term on the dynamics of the full age-structured model. Each point is from the posterior parameter distribution, with the level of NPIs from 1st September onwards varied to generate different underlying growth rates. The decline during the break is the maximum level of NPI at any time for the associated posterior parameter set. (a) shows the temporal reset that captured the time gained by the break; the red line is the theoretical result from the simple theoretical model based on the average rate of decline during the break across all replicates. (b) shows the relative reduction in deaths between 1st October and 1st January.

## 5 Discussion

The pandemic of SARS-CoV-2, and the number of severe cases of COVID-19 worldwide, has necessitated the adoption of a range of control measures in different countries. In the absence of a vaccine or other medical approaches, control has had to rely on very disruptive non-pharmaceutical interventions. In the UK, the implementation of such NPIs during the lockdown period (23rd March - 13th May) reversed the nationally increasing hospitalisations and deaths (*r* ≈ 0.2) and led to a steady decline (*r* ≈ −0.05) [4, 5]. The gradual relaxing of NPIs has generated a slowly increasing growth rate, which we estimate became positive again (leading to exponential growth) in mid-August [7]. Against this backdrop of rising cases, we have investigated the benefits of a planned 2-week precautionary break (or ‘circuit-break’), where strict rules are reapplied in an attempt to drive down cases before their imposition becomes necessary to prevent health system overload. Such breaks are not in themselves long-term solutions, but may allow other methods that work best with low numbers of cases (such as test-trace-and-isolate) to reassert control and they may be more sustainable than other NPI imposition approaches.

Both simple analytical approaches and an age-structured model fit to a range of UK data show that planned precautionary break could be highly effective short-term control measures. Compared to an absence of such controls, precautionary breaks generate a reduction in infection, hospitalisations and deaths, which is most pronounced when the growth rate is moderately high (doubling time of more than 10 days). This reduction in infection can be interpreted as a temporal reset, taking the level of infection back (in time) to a lower value, allowing greater opportunity for additional public health measures to be enacted or take effect.

Unsurprisingly, breaks that generate the most stringent application of NPIs are associated with the greatest immediate rates of declines and, as a consequence, the greatest public health benefits. Therefore, the success of any action is highly contingent on the adherence of the populous to the recommendations and rules; a break is only effective if there is an average increase in level of social distancing across the country. We also consistently find that the optimal time for a break is always now; there are no good epidemiological reasons to delay the break as this will simply push back any benefits until later, leaving more time for additional cases to accumulate. We have focused on combining the break with the school half-term holiday in October, but the same logic would apply to the Christmas holidays (perhaps extending them for a week into 2021) or to the spring half-term.

Ultimately, such short-term precautionary breaks will only be effective if (i) their planned nature helps to minimise the associated societal harms and economic losses, (ii) there is good compliance with the measures across all regions and sections of society, and (iii) the reduction in cases is used to regain control and bring the growth rate back below zero.

## Data Availability

Data on cases were obtained from the COVID-19 Hospitalisation in England Surveillance System (CHESS) data set that collects detailed data on patients infected with COVID-19. Data on COVID-19 deaths were obtained from Public Health England. These data contain confidential information, with public data deposition non-permissible for socioeconomic reasons. The CHESS data resides with the National Health Service (www.nhs.gov.uk) whilst the death data are available from Public Health England (www.phe.gov.uk).

## Author contributions

**Conceptualisation:** Matt J. Keeling; Graham F. Medley.

**Data curation:** Matt J. Keeling; Glen Guyver-Fletcher; Alexander Holmes.

**Formal analysis:** Matt J. Keeling.

**Investigation:** Matt J. Keeling.

**Methodology:** Matt J. Keeling.

**Software:** Matt J. Keeling; Edward M. Hill; Louise Dyson; Michael J. Tildesley.

**Validation:** Matt J. Keeling; Edward M. Hill; Louise Dyson; Michael J. Tildesley.

**Visualisation:** Matt J. Keeling.

**Writing - original draft:** Matt J. Keeling; Graham F. Medley; Edward M. Hill; Michael J. Tildesley; Louise Dyson.

**Writing - review & editing:** Matt J. Keeling; Graham F. Medley; Edward M. Hill; Glen Guyver-Fletcher; Alexander Holmes; Louise Dyson; Michael J. Tildesley.

## Patient and public involvement

This was an urgent public health research study in response to a Public Health Emergency of International Concern. Patients or the public were not involved in the design, conduct, or reporting of this rapid response research.

## Ethical considerations

Data from the CHESS database were supplied after anonymisation under strict data protection protocols agreed between the University of Warwick and Public Health England. The ethics of the use of these data for these purposes was agreed by Public Health England with the Government’s SPI-M(O) / SAGE committees.

## References

[1] World Health Organization. Novel Coronavirus (2019-nCoV) Weekly Epidemiological Update. (2020). URL https://www.who.int/docs/default-source/coronaviruse/situation-reports/ 20201005-weekly-epi-update-8.pdf. [Online] (Accessed: 06 October 2020).

[2] Hale T, Petherick A, Phillips T, Webster S. Variation in government responses to covid-19. Blavatnik school of government working paper 31 (2020).

[3] Rowthorn R, Maciejowski J. A cost–benefit analysis of the covid-19 disease. Oxford Review of Economic Policy 36(Supplement 1):S38–S55 (2020).

[4] Flaxman S, Mishra S, Gandy A, Unwin HJT, Mellan TA, et al. Estimating the effects of non-pharmaceutical interventions on covid-19 in europe. Nature 584(7820):257–261 (2020).

[5] Davies NG, Kucharski AJ, Eggo RM, Gimma A, Edmunds WJ, et al. Effects of non-pharmaceutical interventions on covid-19 cases, deaths, and demand for hospital services in the uk: a modelling study. The Lancet Public Health (2020).

[6] Riley S, Ainslie KE, Eales O, Walters CE, Wang H, et al. Resurgence of sars-cov-2 in england: detection by community antigen surveillance. medRxiv (2020).

[7] Anderson RM, Hollingsworth TD, Baggaley RF, Maddren R, Vegvari C. Covid-19 spread in the uk: the end of the beginning?The Lancet 396(10251):587–590 (2020).

[8] Riley S, Ainslie KE, Eales O, Walters CE, Wang H, et al. High prevalence of sars-cov-2 swab positivity in england during september 2020: interim report of round 5 of react-1 study. medRxiv (2020).

[9] Pierce M, Hope H, Ford T, Hatch S, Hotopf M, et al. Mental health before and during the covid-19 pandemic: a longitudinal probability sample survey of the uk population. The Lancet Psychiatry (2020).

[10] Office for National Statistics. Coronavirus and the impact on output in the UK economy: August 2020 (2020). URL https://www.ons.gov.uk/economy/grossdomesticproductgdp/articles/coronavirusandtheimpactonoutputintheukeconomy/august2020. [Online] (Accessed: 10 October 2020).

[11] Davis EL, Lucas TC, Borlase A, Pollington TM, Abbott S, et al. An imperfect tool: Covid-19’test & trace’success relies on minimising the impact of false negatives and continuation of physical distancing. medRxiv (2020).

[12] Cooper B, Medley G, Stone S, Kibbler C, Cookson B, et al. Methicillin-resistant staphylococcus aureus in hospitals and the community: stealth dynamics and control catastrophes. Proceedings of the National Academy of Sciences 101(27):10223–10228 (2004).

[13] Kretzschmar ME, Rozhnova G, Bootsma MC, van Boven M, van de Wijgert JH, et al. Impact of delays on effectiveness of contact tracing strategies for covid-19: a modelling study. The Lancet Public Health 5(8):e452–e459 (2020).

[14] Keeling MJ, Hill E, Gorsich E, Penman B, Guyver-Fletcher G, et al. Predictions of COVID-19 dynamics in the UK: short-term forecasting and analysis of potential exit strategies. medRxiv page 2020.05.10.20083683 (2020). doi:10.1101/2020.05.10.20083683.

[15] Keeling MJ, Dyson L, Guyver-Fletcher G, Holmes A, Semple MG, et al. Fitting to the UK COVID-19 outbreak, short-term forecasts and estimating the reproductive number. medRxiv page 2020.08.04.20163782 (2020). doi:10.1101/2020.08.04.20163782.

